# Long-term Outcomes of Peripheral Artery Disease In Veterans: Analysis of the PEripheral ARtery Disease Long-term Survival Study (PEARLS)

**DOI:** 10.1101/2024.08.20.24312328

**Authors:** Saket Girotra, Qiang Li, Mary Vaughan-Sarrazin, Brian C. Lund, Mohammad Al-Garadi, Joshua A. Beckman, Rohit Nathani, Richard M. Hoffman, Paul S. Chan, Subhash Banerjee, Shirling Tsai, Dharam J. Kumbhani, Nicole Minniefield-Young, Kim G. Smolderen, Shipra Arya, Cathy Nguyen, Michael E. Matheny, Glenn T. Gobbel

**Affiliations:** University of Texas Southwestern Medical Center (SG, QL, JAB, RN, NMY, DJK, ST, CN), North Texas Veterans Affairs Medical Center (SG, QL, NMY, ST, CN) & Baylor Scott & White Hospital (SB), Dallas, TX; University of Iowa Carver College of Medicine (MVS, BCL, RMH) and Iowa City Veterans Affairs Medical Center (MVS, BCL), Iowa City, IA; Vanderbilt University Medical Center (MAG, GTG, MEM) and Tennesse Valley Health System Veterans Affairs Medical Center (MAG, GTG, MEM), University of Missouri-Kansas City (PSC) & Saint Luke’s Mid America Heart Institute (PSC), Kansas City, MO; Yale School of Medicine (KGS), Stanford School of Medicine (SA) & Palo Alto Veterans Affairs Medical Center (SA), Palo Alto, CA

**Author notes:** Address for Correspondence: Saket Girotra MD, SM Associate Professor of Medicine University of Texas Southwestern Medical Center 5323 Harry Hines Blvd, Suite E5.723 Dallas, TX 75390.

## Abstract

**Background:** Contemporary research in peripheral artery disease (PAD) remains limited due to lack of a national registry and low accuracy of diagnosis codes to identify PAD patients in electronic health records.

**Methods & Results:** Leveraging a novel natural language processing (NLP) system that identifies PAD with high accuracy using ankle brachial index (ABI) and toe-brachial index (TBI) values, we created a registry of 103,748 patients with new onset PAD patients in the Veterans Health Administration (VHA). Study endpoints include mortality, cardiovascular (hospitalization for acute myocardial infarction or stroke) and limb events (hospitalization for critical limb ischemia or major amputation) and were identified using VA and non-VA encounters. The mean age was 70.6 years; 97.3% were males, and 18.5% self-identified as Black race. The mean ABI value was 0.78 (SD: 0.26) and the mean TBI value was 0.51 (SD: 0.19). Nearly one-third (32.4%) patients were currently smoking and 35.4% formerly smoked. Prevalence of hypertension (86.6%), heart failure (22.7%), diabetes (54.8%), renal failure (23.6%), and chronic obstructive pulmonary disease (35.4%) was high. At 1-year, 9.4% of patients had died. The 1-year incidence of cardiovascular events was 5.6 per 100 patient-years and limb events was 4.5 per 100 patient-years.

**Conclusions:** We have successfully launched a registry of >100,000 patients with a new diagnosis of PAD in the VHA, the largest integrated health system in the U.S. The ncidence of death and clinical events in our cohort is high. Ongoing studies will yield important insights regarding improving care and outcomes in this high-risk group.

## Introduction

An estimated 8 to 12 million Americans suffer from lower extremity peripheral artery disease (PAD).^1,2^ Although PAD shares risk factors (e.g., diabetes, smoking, hypertension) with other atherosclerotic vascular diseases such as coronary artery disease (CAD), patients with PAD experience a higher rate of cardiovascular (myocardial infarction, stroke) and limb events (critical limb ischemia, amputation), which contribute to excess disability, death and medical expenditures.^3–6^ Importantly, racial and ethnic disparities in the incidence and outcomes of PAD have been reported, with a higher incidence of lower extremity amputations in Black, as compared with White, individuals with PAD.^7,8^

While clinical practice guidelines recommend a multi-pronged strategy including medications (e.g., statins), risk factor control (blood pressure [BP] and diabetes), lifestyle changes, and revascularization in select patients,^9^ the above therapies are unevenly applied in clinical practice.^10–16^ This may be due to a paucity of contemporary evidence regarding existing treatments on long-term outcomes. Unlike CAD, there are also unique challenges to reliably identifying PAD patients within electronic health records (EHR) data; existing PAD diagnosis codes historically have had low positive predictive values ranging from 27.5% to 69.4%.^17–20^ The limited accuracy of diagnosis codes have hampered the development of large registries to improve care and outcomes in PAD.

We recently developed and validated a natural language processing (NLP) system that can extract ankle-brachial index (ABI) and toe-brachial index (TBI) values from ABI test reports documents in the Veterans Affairs (VA) EHR and use these direct measures of ABI and TBI values to identify patients with PAD.^21^ In contrast to using only International Classification of Diseases (ICD) codes, our NLP-based approach had a sensitivity of 89.3%, specificity of 92.3%, and positive predictive value of 92.3% for identifying PAD.^21^ Leveraging this NLP system, we have launched the **<u>Pe</u>**ripheral **<u>AR</u>**tery **<u>L</u>**ong-term **<u>S</u>**urvival Study (PEARLS), a longitudinal study of PAD in the Veterans Health Administration (VHA) – the largest integrated health system in the U.S. In this paper, we describe the creation of the PEARLS registry – a contemporary cohort of newly diagnosed PAD patients identified using our novel NLP system in the VHA EHR. We also provide descriptive data on the baseline characteristics of our cohort including data on demographics, co-morbidities, medications, and longitudinal data on 1-year incidence of mortality, cardiovascular and limb events.

## Methods

This work was approved by the Institutional Review Board (IRB) and Research & Development Committees at Dallas, Iowa City and Tennessee Valley Health System VA centers as well as the IRBs at University of Texas Southwestern Medical Center and University of Iowa.

The primary objective of the study was to develop a longitudinal cohort of patients with incident PAD using our novel NLP system. In the first stage of cohort creation, we executed the NLP system on the entire corpus of ABI report documents in the VA’s EHR. Among patients identified as having PAD, we applied additional criteria to restrict our cohort to patients with a new diagnosis of PAD and those who use VA regularly for healthcare **(Figure 1)**. Next, we identified baseline demographics, co-morbidities, smoking status, medications, vital signs, and laboratory values for our cohort. We followed the cohort longitudinally for clinical events using a combination of VA and non-VA (Medicare) data sources to ensure complete follow up.

**Figure 1.**
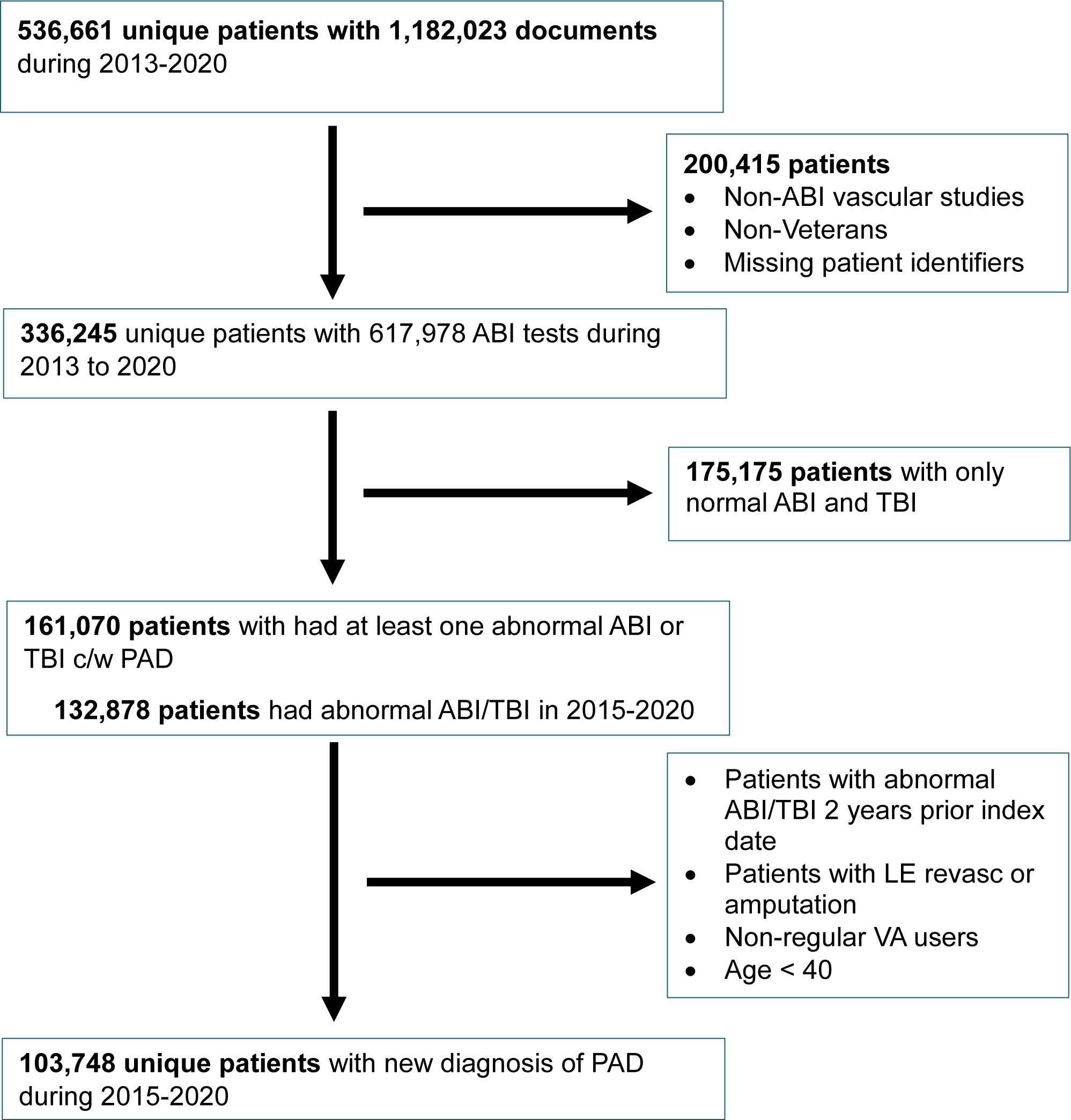
Study Flowchart

### Data Sources

The list of data sources used in the current study are summarized in **Table S1.** The VA Corporate Data Warehouse (CDW) is a national repository of clinical data for patients receiving care at a VA facility. Because the VA is a single-payer health care system, it generates data from clinical functions (e.g. clinical notes, procedural and radiology reports) and claims-based functions (e.g. clinical diagnoses and procedures using International Classification of Diseases Ninth and Tenth – Clinical Modification (ICD-9 CM and ICD-10 CM) and Current Procedural Terminology [CPT] codes). The above data are available for inpatient and outpatient encounters. In addition, data on smoking status, vital signs from each clinical encounter, laboratory test results and prescriptions filled at a VA pharmacy are also available. **Table S1** includes details of data domains within the CDW that were used for identifying study variables. Unique patient identifiers allow linkage of data across domains to create a comprehensive patient record.

Since Veterans may also receive care at a non-VA facility, relying on VA data alone may not provide complete information regarding clinical events after PAD diagnosis. Therefore, we obtained two additional sources of data to supplement clinical data available in the VHA. These include the a) Fee-basis files and Consolidated Dataset (CDS) files and b) Medicare claims. The Fee-basis (2019 and earlier) and CDS (2020 onwards) data include claims arising from services provided to Veterans at a non-VA facility (or by a non-VA provider) that are paid by the VA. Typically, the VA pays for emergent care received by a Veteran at a non-VA facility (e.g., hospitalization for acute myocardial infarction (MI) or stroke). The VA also pays for routine visits at a non-VA facility if travelling to a VA facility for the same services imposes undue burden (e.g, vascular surgery clinic at a non-VA facility near a Veteran’s home if comparable services at a VA facility require long travel). Each year the VA Information Resource Center (VIReC) obtains 100% Medicare enrollment and claims data from the Center for Medicare and Medicaid (CMS) for all Veterans. These include Part A (fee for service), Part B & Carrier (outpatient), Part C (Advantage), and Part D (medications). For our cohort, we obtained all Part A (fee-for-service), Part B & Carrier (outpatient facility & physician claims), Part C (Medicare Advantage) and Part D (medications) claims for our cohort during 2013-2022 from VIReC. Part C data were only available through 2021. Because >70% of Veterans in our cohort were older than 65 years of age (Medicare eligible) at the time of PAD diagnosis, the addition of Medicare claims ensured more comprehensive follow up from clinical encounters outside the VA especially those older than 65 years of age. Thus, the availability of these additional data sources ensures near complete follow up of our cohort of PAD patients.

### ABI Reports

Within the CDW data, we identified all reports of ABI tests performed during 2013-2020 at a VA facility. Depending on the facility, ABI tests maybe performed by vascular surgery, radiology or primary care. Therefore, ABI report documents can be found in either the Text Integration Utility (TIU) or Radiology domain within Unstructured Clinical data in CDW. In both domains, documents are organized into document types with each document type having a unique ID and an associated title (e.g., “Vascular Lab ABI report”). The document type IDs are unique to each VA facility and can be used to identify all individual documents within that facility (e.g., all ABI test results that are reported using the “Vascular Lab ABI report” title would have a common document type ID). Often, a facility uses more than one document type for reporting ABI results especially if ABI tests are performed by multiple specialties at that facility. We included all possible document type IDs used for reporting ABI results at each facility. Finally, in some instances, a facility used a ‘generic’ document type ID (e.g. “Vascular Lab Report”) for reporting both ABI studies and non-ABI vascular studies (e.g., carotid duplex, etc.). Such generic document type IDs were also included. In our validation study, the NLP system extracted no false positive ABI or TBI values from non-ABI vascular studies.^21^ A total of 424 document type IDs used for reporting ABI results in the VHA during 2013-2020 were identified which corresponded to 1,182,023 individual documents.

### NLP Deployment

Since the NLP system was developed using only a small random sample of individual ABI documents (800 total), certain adaptations were necessary to handle the large document corpus without losing accuracy. In our development work, we used the same text-based features for random forest model training and subsequent testing. However, this approach proved to be infeasible at scale due to temporal and spatial requirements, which requires re-training the random forest model for every document. To overcome this challenge, we first redesigned and refactored the code underlying the tool and re-tested model performance using the previous training and test corpora to ensure no change in performance. We also built a disk-based storage system that could hold millions of files and allow for rapid retrieval of documents without consuming excessive disk space. Finally, we created a database programming script that can identify PAD based on extracted ABI and TBI values (**Figure S1**). With these changes, we were able to process all of the documents in a total of 95 hours. This equated to 3.48 documents processed per second or over 300,000 documents per day.

### Creation of the PEARLS Cohort

Since our overall objective was to identify Veterans with a <u>new diagnosis</u> of PAD during 2015 to 2020, we intentionally added two additional years of ABI data (2013-2014) as a ‘look-back’ period. This was done to ensure that patients identified as PAD based on abnormal ABI or TBI for the first time in 2015-2020 did not also have abnormal ABI or TBI value during the prior 2 years.

**Figure 1** shows the derivation of our study cohort. After processing ∼1.2 million documents and excluding non-ABI vascular studies, non-Veterans and patients missing unique IDs, a total of 336,245 patients with 617,978 ABI report documents remained. Among these patients, 161,070 (47.9%) met criteria for PAD (i.e., ABI value of ≤0.9 or a TBI value ≤0.7, **Figure S1**). For each patient, all ABI reports were arranged in chronological order, and the index date (date of PAD diagnosis) was defined as the earliest date of abnormal ABI or TBI value in any limb. From this sample, we excluded patients with evidence of PAD in the 2 years prior to 2015 or a history of lower extremity revascularization or amputation in the 2 years before the index date. Additional exclusions were non-regular use of VA services (less than 2 outpatient, or 1 inpatient and 1 outpatient visit in the prior 2 years), age less than 40 years on index date (as PAD is rare in those younger than 40 years), and death before the index date (data coding errors). Our final sample included 103,748 patients with a new diagnosis of PAD during 2015 to 2020.

### Study Outcomes

The registry collected a number of outcomes relevant to PAD. All-cause mortality was determined using the VA Death Ascertainment file. Other important outcomes that were collected included cardiovascular events (hospitalization for acute myocardial infarction [MI]) or ischemic stroke) and limb events (CLI and major amputation). Major amputation was defined as any amputation above the ankle joint. The diagnosis and procedure codes used to identify these events are listed in **Table S2.** Hospitalization for MI, stroke, and CLI were included only when they were reported as primary diagnosis on the inpatient encounter as multiple prior studies have demonstrated better accuracy only when the primary diagnosis is considered. ^22–26^

### Study Variables

Demographic characteristics included age, sex, race (White, Black, other), and ethnicity (Hispanic vs. non-Hispanic). Smoking status up to 12 months before the index date was determined using the health factors table in VHA CDW, which contains details on routine screenings and social factors that impact health. Co-morbidities were based on clinical diagnosis entered in inpatient and outpatient encounters from the VA, Fee-basis and CDS files during the 2 years prior to the index date, and defined using algorithms originally developed by Elixhauser, and updated by Quan.^27^ We used *Vital Signs* data from outpatient encounters to determine baseline systolic and diastolic blood pressure during 24 months before the index date. We used *Laboratory* data to determine baseline levels of total cholesterol, low-density lipoprotein (LDL), high-density lipoprotein (HDL), triglycerides and hemoglobin A1c levels within 12 months before the index date. We also included the ABI and TBI values extracted by the NLP system from the qualifying ABI report. We identified prescription fills using VA Pharmacy and Medicare Part D data to determine baseline use of lipid lowering drugs, antiplatelet drugs, anticoagulants, anti-hypertensive and anti-diabetic drugs. For each drug, we defined active use at baseline as filling a prescription for that drug prior to the index date within a time-period that was less than or equal to twice the days’ supply dispensed. For example, a PAD patient with an index date of August 1, 2015 who filled a 30-day supply of atorvastatin on June 15, 2015 (46 days before the index date) would be considered an active statin user as the drug was filled within 60 days (2×30 days’ supply).

### Statistical Analyses

Baseline characteristics of our study cohort were reported using means (standard deviations) and frequencies (proportions) as appropriate. Next, we examined the overall rate of each study endpoint: mortality, hospitalization for MI or stroke (cardiovascular events) and CLI and amputation (limb events) at 1-year follow-up. We also constructed survival curves as well as the cumulative index function for the secondary endpoints over a 1-year follow-up period. For non-fatal endpoints, the incidence rates were computed after accounting for the competing risk of death and expressed per 100 patient-years. Finally, for non-fatal endpoints, we also quantified the proportion of events that were identified using different data sources (VA data, Fee-basis, Medicare fee-for-service, and Medicare Advantage).All analyses were performed using SAS version 9.4 (SAS Institute, Cary, North Carolina).

## Results

**Table 1** shows baseline characteristics of our cohort. Among 103,748 eligible patients with PAD, the mean age was 70.6 years (standard deviation [SD] of 9.1 years), and the majority (97.3%) were men. Nearly 1 in 5 patients (18.5%, N=19,208) were of Black race and 3.0% (N=3,130) were of Hispanic ethnicity. The mean (SD) ABI and TBI in the worst limb was 0.78 (0.26) and 0.51 (0.19), respectively. Among 84% of patients with available data on smoking within 1 year of index data, 32.2% were currently smoking and 35.7% formerly smoked. Overall, there was a high prevalence of hypertension (86.6%), coronary artery disease (42.9%), cerebrovascular disease (23.8%), diabetes (54.8%), chronic kidney disease (23.6%) and chronic obstructive pulmonary disease (35.4%). At baseline, the mean systolic blood pressure was 133 mm Hg, and mean diastolic blood pressure was 71 mm Hg, and 58.4% (N=98,869 with available BP data) had a systolic blood pressure >130 mm Hg or diastolic blood pressure >80 mm Hg. The mean values of cholesterol, triglycerides and hemoglobin A1c are also included in **Table 1**.

**Table 1.** Baseline Characteristics of Veterans with New Diagnosis of PAD 2015-2020

| **Variable** | **N=103,748** |
| --- | --- |
| **Demographics** |  |
| Mean age, years (SD) | 70.6 (9.1) |
| Male sex | 100,972 (97.3%) |
| Race |  |
| White | 77,321 (74.5%) |
| Black | 19,208 (18.5%) |
| Other | 1,660 (1.6%) |
| Multiple races | 723 (0.7%) |
| Unknown | 4836 (4.7%) |
| Ethnicity |  |
| Hispanic | 3,130 (3.0%) |
| Not Hispanic | 96,951 (93.4%) |
| **Smoking Status\*** |  |
| Current | 28,086 (32.2%) |
| Former | 31,132 (35.7%) |
| Non-smoker | 13,160 (15.1%) |
| Unknown | 14,906 (17.1%) |
| **Co-morbidities** |  |
| Hypertension | 89,827 (86.6%) |
| Diabetes | 56,904 (54.8%) |
| Chronic kidney disease | 24,467 (23.6%) |
| COPD | 36,676 (35.4%) |
| Arrhythmia | 31,390 (30.3%) |
| Heart failure | 23,548 (22.7%) |
| Coronary artery disease | 44,462 (42.9%) |
| Prior history of MI | 11,252 (10.8%) |
| Cerebrovascular disease | 24,731 (23.8%) |
| Cancer | 16,285 (15.7%) |
| Dementia | 5,232 (5.0%) |
| Depression | 31,750 (30.6%) |
| Chronic liver disease | 9,699 (9.3%) |
| Obesity | 25,244 (24.3%) |
| Anemia | 11,401 (11.0%) |
| Valvular heart disease | 11,010 (10.6%) |
| Weight loss | 7,814 (7.5%) |
| **Blood Pressure, Mean (SD)\*** |  |
| Systolic | 133 (19) |
| Diastolic | 73 (11) |
| **Laboratory Values, Mean (SD)\*** |  |
| Total Cholesterol | 162mg/dL (44.6) |
| LDL-Cholesterol | 91.0mg/dL (37.2) |
| HDL-Cholesterol | 43.1mg/dL (14.1) |
| Triglycerides | 157.1mg/dL (123.6) |
| Hemoglobin A1c | 6.9% (1.6) |
| **ABI and TBI Values, Mean (SD)\*** |  |
| ABI in worst limb | 0.78 (0.26) |
| TBI in worst limb | 0.51 (0.19) |

Data in the table represent N(%), unless otherwise specified
\*Data on smoking status were available in 87,284 (84.1%), vital signs in 98,869 (95.3%) patients; patients, total cholesterol in 89,818 (86.6%) patients, LDL-cholesterol in 89,117 (85.9%) patients, HDL-cholesterol in 89,569 (86.3%) patients, triglycerides in 88,905 (85.6%) patients and hemoglobin A1c in 79,918 (77.0%) patients, ABI values in 99,266 (95.7%) patients, and TBI values in 57,010 (54.5%) patients

**Table 2** shows baseline use of medications in our PAD cohort. Nearly two-thirds (65.4%) of PAD patients were on a statin at the time of PAD diagnosis, but only 34.2% were on a high-intensity statin. Among non-statin lipid-lowering medications, use of ezetimibe (1.1%) and Proprotein Convertase Subtilisin/Kexin-9 (PCSK-9) inhibitors (< 0.1%) at baseline was uncommon. Anti-plaletelet drugs were used in 35.0% (asprin: 24.5%, P2Y12 inhibitors: 13.3%, and phosphodiesterase inhibitors: 3.0%) while anticoagulants were used in 12.9% of patients (warfarin: 6.9%, novel oral anticoagulant 6.3%). Only 25 patients (<0.1%) were taking low dose rivaroxaban at baseline. Overall, 81.2% patients were receiving an anti-hypertensive with angiotensin converting enzyme-inhibitors, angiotensin receptor blockers and angiotensin receptor neprilysin inhibitor being the most common medications. Among the 41.3% of patients receiving anti-diabetic agents, metformin was the most common agent (24.0% patients) followed by sulfonylureas and insulin. Baseline use of glucagon-like peptide (GLP-1) agonists and sodium glucose transporter-2 (SGLT-2) inhibitors was low.

**Table 2.** Medications at Baseline in PAD Cohort

|  |  |
| --- | --- |
|  | **N=103,748**<br>n (%) |
| ***Lipid Lowering Therapy*** | **68,337 (65.9)** |
| Statins | 67,858 (65.4) |
| High-intensity statin | 35,499 (34.2) |
| Ezetimibe | 1,141 (1.1) |
| PCSK-9 | 10 (0.0) |
| ***Antiplatelet agents\**** | **36,312 (35.0)** |
| Aspirin | 25,449 (24.5) |
| P2Y12 Inhibitor | 13,749 (13.3) |
| Phosphodiesterase inhibitor | 3,149 (3.0) |
| ***Anticoagulants*** | **13,410 (12.9)** |
| Warfarin | 7,116 (6.9) |
| NOAC | 6,501 (6.3) |
| ***Anti-hypertensive agents*** | **84,226 (81.2)** |
| ACE-I/ARB/ARNI | 57,517 (55.4) |
| Beta blockers | 49,484 (47.7) |
| Diuretics | 42,963 (41.1) |
| Calcium channel blocker | 31,708 (30.6) |
| Vasodilators | 3,758 (3.6) |
| Alpha blocker | 8,728 (8.4) |
| Central acting | 1,887 (1.8) |
| ***Anti-diabetic agents*** | **42,826 (41.3)** |
| Metformin | 24,931 (24.0) |
| Insulin | 22,952 (22.1) |
| DPP-4 inhibitors | 3,121 (3.0) |
| GLP-1 agonists | 1,441 (1.4) |
| SGLT-2 inhibitors | 1,430 (1.4) |
| Thiazolidinediones | 1,187 (1.1) |
| Alpha-glucosidase inhibitors | 442 (0.4) |
| Meglitinides | 97 (0.1) |

Data in the table represent number (%).
**Abbreviations:** ACE-I: Angiotensin Converting Enzyme-Inhibitor; ARB: Angiotensin Receptor Blocker; ARNI: Angiotensin Neprilysin Inhibitor; DPP-4: Dipeptidyl Peptidase-4; GLP-1: Glucagon-like peptides; NOAC: Novel oral anticoagulant; PCSK-9: Proprotein convertase subtilin/kexin type 9; SGLT-2: sodium glucose transporter-2

The number of clinical events and the incidence of study endpoints are reported in **Table 3** and **Figures 2 & Figure 3**, respectively. Overall, 9,723 participants (9.4%) died within 1-year of the index date. During 1-year follow-up, 2,706 (2.6%) experienced an acute MI and 2881 (2.8%) experienced a stroke, 2465 (2.4%) experienced a CLI hospitalization, and 2616 (2.5%) underwent a major amputation. After accounting for the competing risk of mortality, the 1-year incidence rate of cardiovascular events was 5.6 per 100 patient-years (**Figure 3A**), which included acute MI (2.8 per-100 patient-years) and stroke (3.0 per-100 patient-years). The 1-year incidence of limb events was 4.5 per-100 patient-years **(Figure 3B)** which comprised of hospitalization for CLI (2.5 per-100 patient-years) and major amputation (2.7 per-100 patient-years).

**Table 3.**
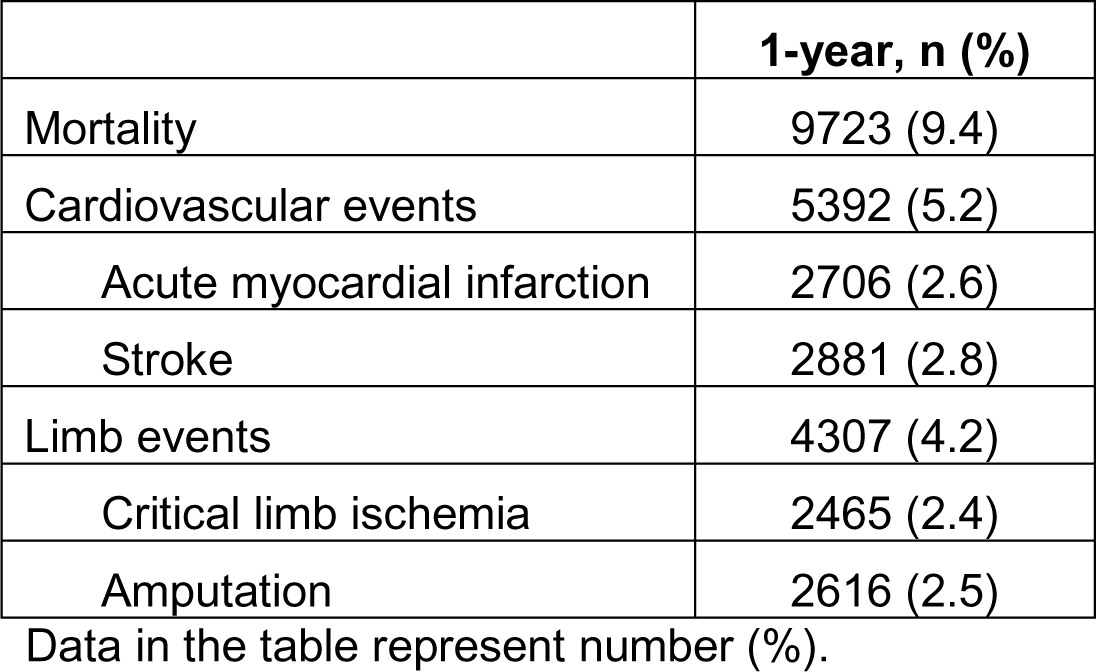
The number of patients who experienced a study endpoint at 1-year

|  | **1-year, n (%)** |
| --- | --- |
| Mortality | 9723 (9.4) |
| Cardiovascular events | 5392 (5.2) |
| Acute myocardial infarction | 2706 (2.6) |
| Stroke | 2881 (2.8) |
| Limb events | 4307 (4.2) |
| Critical limb ischemia | 2465 (2.4) |
| Amputation | 2616 (2.5) |

Data in the table represent number (%).

**Figure 2.**
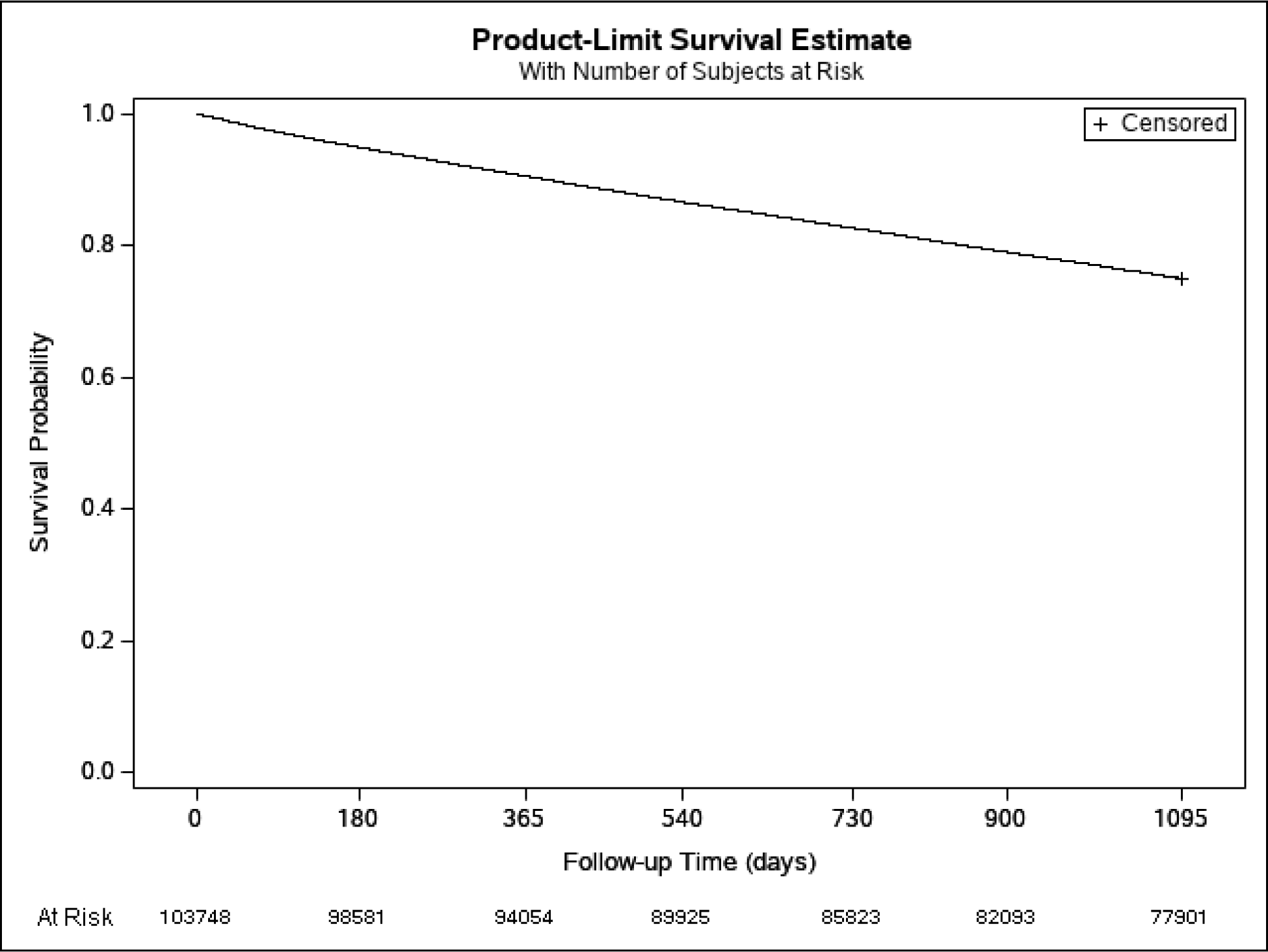
Kaplan Meier Curve for Survival at 1-year. The figure shows the Kaplan-Meier estimates of survival over 1-year follow up

**Figure 3.**
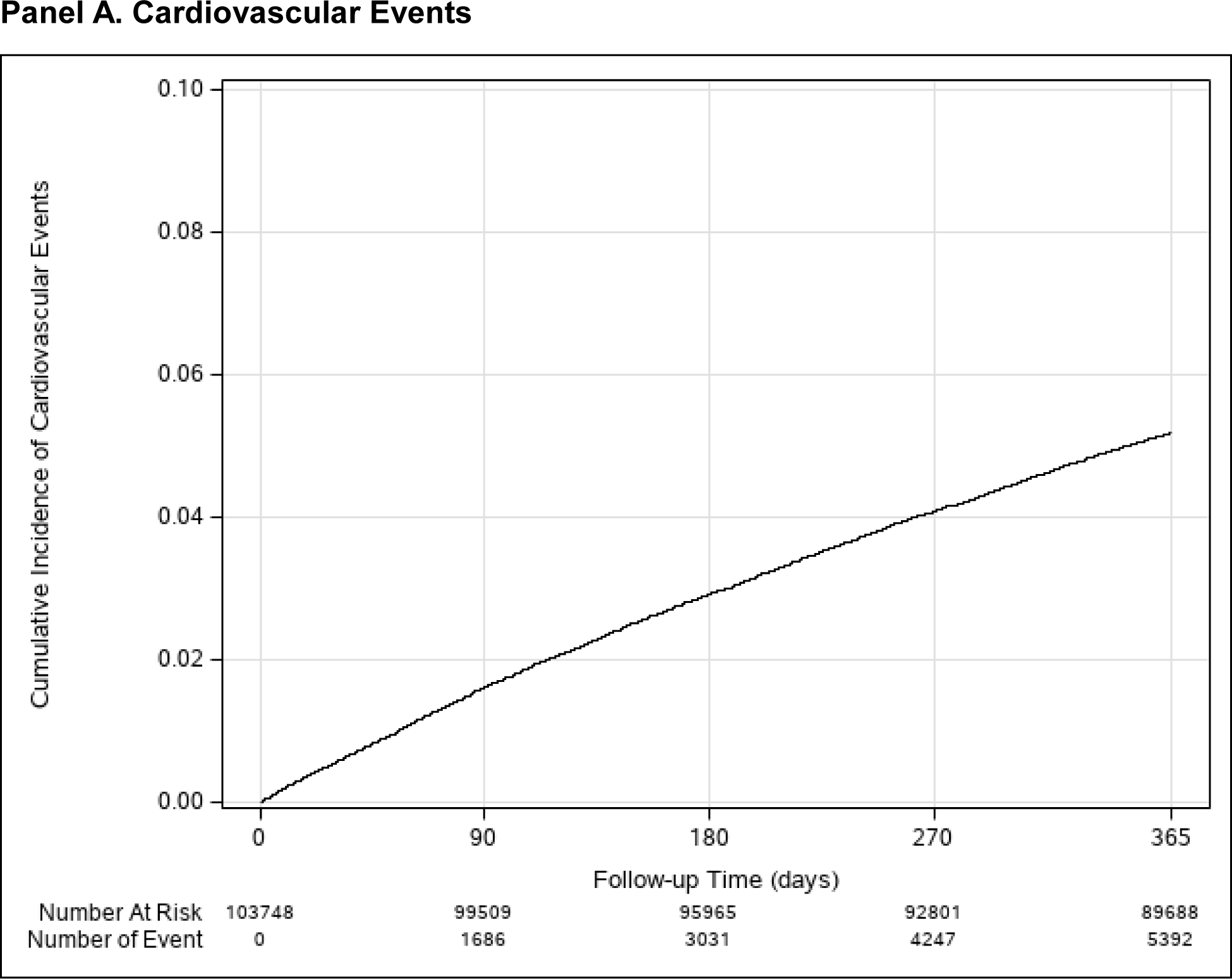
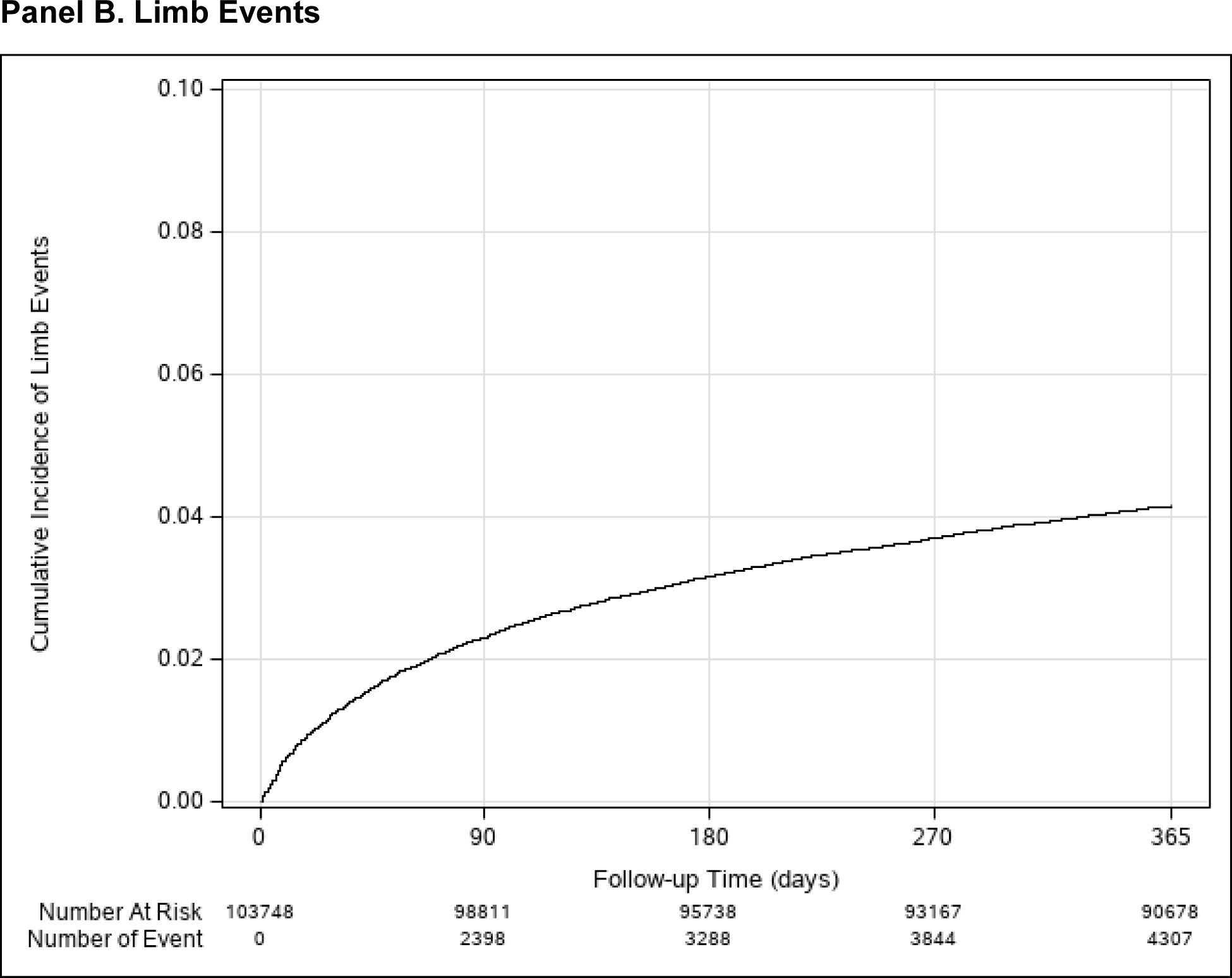
Cumulative Incidence of Cardiovascular Events and Limb Events. Panel A shows the cumulative incidence of cardiovascular events (hospitalization for acute myocardial infarction or stroke) through 1-year. Panel B shows the cumulative incidence of limb events (hospitalization for critical limb ischemia or amputation). The cumulative incidence function curves were estimated after accounting for competing risk of mortality.

**Table S3** shows the proportion of non-fatal events identified using different data sources. Of the 5329 patients with a cardiovascular event (acute MI or stroke), 1721 (31.9%) were identified in the VHA, 2071 (38.4%) in Fee-basis (non-VA hospital claims paid by the VHA), 1177 (21.8%) and 423 (7.8%) in Medicare fee-for-service and Advantage, respectively. Among 4307 patients with a limb event (CLI or major amputation), 2726 (63.3%) were in the VHA, 660 (15.3%) in Fee-basis, 644 (15.0%) in Medicare fee-for-service, and 277 (6.4%) in Medicare Advantage claims.

## Discussion

Using a previously validated NLP system,^21^ we have created a registry of >100,000 newly diagnosed patients with PAD in the VHA. In contrast to solely using administrative diagnosis and procedure codes that have modest accuracy,^17–20^ identification of PAD in our cohort is based on abnormal ABI and TBI values which are considered the gold standard methods for clinical diagnosis of PAD.^28^ Importantly, our cohort is racially and ethnically diverse with >19,000 individuals self-identified as Black, and 3,000 self-identified as Hispanic. The prevalence of smoking and other co-morbidities, including hypertension, coronary and cerebrovascular disease, diabetes, and chronic kidney disease was high. Rates of all-cause mortality approached 10% at 1-year, with high rates of cardiovascular and limb events, which underscore the enormous impact of PAD in real-world patients.

Prior studies of PAD in large health systems have been limited by the imprecision of using ICD codes for identifying PAD. These studies have reported the sensitivity of ICD codes to range from 34.7% to 76.9% and the positive predictive value to range from 27.5% to 69.4%.^17–20^ The low accuracy of ICD codes for PAD identification likely affects inferences drawn from such studies. To overcome this challenge, we developed an NLP system that can extract ABI and TBI values including laterality from ABI reports in the VHA.^21^ An algorithm based on NLP-extracted ABI and TBI values had a higher sensitivity of 83.3% and positive predictive value of 92.4% as compared ICD diagnosis and procedure codes for identifying PAD.^21^ Building on this prior work, we now report the findings following the implementation of our NLP system in national VA data to develop an inception cohort of newly diagnosed PAD in the VHA during 2015-2020. The PEARLS cohort includes >100,000 Veterans with a new diagnosis of PAD, making it one of the largest contemporary cohorts of PAD to our knowledge. Importantly, through linkage with an array of VA and non-VA data sources, rich information on key variables (e.g., smoking status, co-morbidities, vital signs, laboratory values and medications) as well as comprehensive follow-up is available. Thus, the PEARLS registry is well positioned to evaluate the long-term trajectory of health outcomes, evaluate the association of PAD treatments with long-term outcomes, and identify opportunities for improving care.

There are several advantages of studying PAD in the VHA. First, the VHA provides care to over 9 million Veterans across 130 VA medical centers and 1138 outpatient sites that use a standardized EHR.^29^ Risk factors for PAD which include smoking, hypertension, and diabetes are highly prevalent leading to a high burden of PAD in Veterans. Second, although women are under-represented, our cohort still includes ∼3,000 women representing one of the largest contemporary cohorts of women with PAD. Importantly, racial and ethnic subgroups are well-represented in our study with the inclusion of >19,000 Black individuals and ∼3000 Hispanic individuals, groups that have been under-represented in prior studies. Third, in addition to inpatient and outpatient claims, PEARLS also includes rich information on medications (VA *Pharmacy* and Part D claims), *Vital Signs* captured during clinical encounters, and *Laboratory* results that are generally unavailable in other cohorts. Most importantly, the availability of Fee-basis, CDS and Medicare files ensures comprehensive follow up data for defining clinical exposures (e.g., medications) and clinical endpoints from VA and non-VA encounters (except the small minority of Veterans insured with commercial insurance plans).^30,31^ This overcomes a key limitation of other EHR studies that are often limited in assessing events if they occurred in a different health system. Finally, VA data are updated nightly providing real-time access avoiding delay inherent in studies that use other insurance claims data.

The high incidence of clinical events in our cohort underscores the substantial impact of PAD on patients. Nearly 10% of patients with PAD died within 1 year of diagnosis. The incidence of cardiovascular events was 5.6% per 100 patient-years at 1-year accounting for competing risk of mortality, highlighting that PAD is a marker of malignant vascular phenotype with clinical event rates that far exceed those of patients with stable atherosclerotic cardiovascular disease.^32^ For instance, among 32,961 patients with stable CAD, the incidence of death or cardiovascular events was 7.6% at 5 years.^32^ It was also striking that the incidence of limb events -hospitalization for CLI and major amputation was high as well (4.5 per-100 patient-years) approaching that of cardiovascular events. The downstream impact of a limb event – risk of infection, limited mobility and disability – is enormous and contributes to high mortality and poor quality of life in PAD patients. Thus, our findings support guideline recommendations which consider abnormal ABI as a risk enhancer for primary prevention and the importance of developing strategies for not only evaluating therapies for reducing cardiovascular but also limb events.^33^

The use of an accurate method of identifying PAD patients combined with rich and comprehensive data provides a unique opportunity to enhance contemporary PAD research in important ways. Ongoing studies from our cohort will yield important insights regarding patterns of care delivery for PAD patients (e.g., medications, risk factor control and revascularization) and the extent to which these exposures are associated with long-term outcomes. Moreover, given the sizeable number of Black and Hispanic individuals in our cohort, we will also be able to evaluate whether disparities exist in care and outcomes of PAD patients by race and ethnicity. Because health insurance is not a barrier in the VHA, we will also have a unique opportunity to isolate the contribution of access to healthcare from other social determinants of health in explaining disparities. Finally, the large number of VA sites in our cohort provides an opportunity to understand variation in care patterns across sites with the goal of identifying best practices.

Our study findings should be considered in the context of the following limitations. First, ABIs can be normal in patients with non-compressible vessels. Although we used TBI values to identify PAD in patients with falsely normal ABI values, TBI measurements are not always performed. Thus, patients with non-compressible vessels who did not have TBI measured are not included. Second, data on clinical symptoms and severity (e.g., claudication, rest pain) are also not included. Since routine ABI screening of asymptomatic patients is not universally recommended, we expect that most patients in our cohort have symptomatic PAD. Third, we identified clinical endpoints using a combination of VA claims, non-VA claims paid by the VA or Medicare claims to ensure near complete follow up. However, it is possible that the incidence of clinical endpoints in younger Veterans not insured by Medicare or those with private insurance is underestimated. Fourth, medication data were based on prescriptions that were either filled at a VA pharmacy or using Medicare Part D insurance. ^30,31^ However, the VA provides generous pharmacy benefits for Veterans which include a) modest co-payment not to exceed $11 for all drugs including brand name medications, b) no co-payments for disabled Veterans with service connection and c) the convenience of mail-order and online pharmacy. These incentives likely minimize the potential for missing information on medication exposures in our cohort.^34,35^ Finally, due to its availability over the counter, use of aspirin may be underreported in our study.

## Conclusion

We have successfully developed a registry of >100,000 patients with a new diagnosis of PAD in the Veterans Health Administration using a novel NLP algorithm. Ongoing studies from our cohort will yield important insights into the association between management of PAD and clinical outcomes, with the goal of identifying opportunities for improving care.

**Funding**: The study is funded by the National Heart, Lung and Blood Institute (NHLBI, R01HL166305 and R56HL158803; PI: Girotra; Co-I: Vaughan-Sarrazin, Lund, Al-Garadi, Beckman, Hoffman, Tsai, Smolderen, Arya, Matheny, Gobbel). Dr. Lund receives funding from the VA Office of Research and Development and the VA Office of Rural Health. Dr. Chan and Dr. Girotra also receive receive funding from the NHLBI (R01HL160734) and the American Heart Association. This material is the result of work supported with resources and the use of facilities at the Iowa City, Nashville and North Texas Veterans Affairs. None of the funders had any role in the design and conduct of the study; collection, management, analysis, and interpretation of the data; preparation for publication. The views expressed in this article are those of the authors and do not necessarily reflect the position or policy of the Department of Veterans Affairs or the United States Government.

## Data Availability

Due to the proprietary nature of the data used in the study, data cannot be made available

